# Vaccine effectiveness for preventing COVID-19 hospital admission during pregnancy: a population-based cohort study in England during the Alpha and Delta waves of the SARS-CoV-2 pandemic

**DOI:** 10.1101/2022.09.27.22280397

**Authors:** Matthew L. Bosworth, Ryan Schofield, Daniel Ayoubkhani, Loes Charlton, Vahé Nafilyan, Kamlesh Khunti, Francesco Zaccardi, Clare Gillies, Ashley Akbari, Marian Knight, Rachael Wood, Pia Hardelid, Luisa Zuccolo, Camille Harrison

## Abstract

**Objective:** To estimate vaccine effectiveness (VE) for preventing COVID-19 hospital admission in women first infected with SARS-CoV-2 during pregnancy, and assess how this compares to VE among women of reproductive age who were not pregnant when first infected.

**Design:** Population-based cohort study using national, linked Census and administrative data.

**Setting:** England, United Kingdom, from 8^th^ December 2020 to 31^st^ August 2021.

**Participants:** 815,4777 women aged 18 to 45 years (mean age, 30.4 years) who had documented evidence of a first SARS-CoV-2 infection in NHS Test and Trace data or Hospital Episode Statistics.

**Main outcome measures:** A hospital inpatient episode where COVID-19 was recorded as the primary diagnosis. Cox proportional hazards models, adjusted for calendar time of infection and sociodemographic factors related to vaccine uptake and risk of severe COVID-19, were used to estimate VE as the complement of the hazard ratio for COVID-19 hospital admission.

**Results:** Compared with unvaccinated pregnant women, the adjusted rate of COVID-19 hospital admission was 76% (95% confidence interval 69% to 82%) lower for single-vaccinated pregnant women and 83% (75% to 88%) lower for double-vaccinated pregnant women. These estimates were similar to those found for non-pregnant women: 79% (76% to 81%) for single-vaccinated and 82% (80% to 83%) for double-vaccinated. Among those vaccinated more than 90 days before infection, being double-vaccinated was associated with a greater reduction in risk than being single-vaccinated.

**Conclusions:** COVID-19 vaccination is associated with reduced rates of severe illness in pregnant women infected with SARS-CoV-2, and the reduction in risk is similar to that for non-pregnant women. Waning of vaccine effectiveness occurs more quickly after one dose of a vaccine than two doses.

**What is already known on this topic:** Being pregnant is a risk factor for severe illness and mortality following infection with SARS-CoV-2.

Existing evidence suggests that COVID-19 vaccines are effective for preventing severe outcomes in pregnant women.

However, research directly comparing vaccine effectiveness between pregnant and non-pregnant women of reproductive age at the population level are lacking.

**What this study adds:** Our study provides real-world evidence that COVID-19 vaccination reduces the risk of hospital admission by a similar amount for both women infected with SARS-CoV-2 during pregnancy and women who were not pregnant when infected, during the Alpha and Delta dominant periods in England.

## Introduction

Physiological changes that take place during pregnancy (e.g., insulin resistance, low blood pressure, and changes to respiration) place pregnant women at elevated risk of experiencing severe outcomes of coronavirus disease 2019 (COVID-19). [1] While the absolute risk of being admitted to hospital or dying with COVID-19 during pregnancy is low, [2, 3, 4] COVID-19 in pregnancy is associated with maternal and perinatal morbidity and mortality. [5, 6, 7] A meta-analysis of 21 studies reported that pregnant and postpartum women with COVID-19 are at increased risk of admission to intensive care and all-cause mortality compared with pregnant and postpartum women without COVID-19. [8]

COVID-19 vaccines have been demonstrated to be highly effective at reducing the risk of COVID-19 hospitalisation and death in both clinical trials and real-world observational studies. [9, 10, 11] Although pregnant women were not included in the original trials, a meta-analysis of three observational studies found that two doses of a mRNA vaccine was 89.5% effective at preventing SARS-CoV-2 infection during pregnancy. [12] Other studies have shown that vaccination reduces the risk of severe illness in pregnant women infected with SARS-CoV-2. [13, 14] Consistent with these observations, the majority of pregnant women admitted to hospital or intensive care units for COVID-19 in the UK and across Europe were unvaccinated. [15, 16, 17] Despite accumulating evidence for efficacy and safety of COVID-19 vaccines for pregnant women, vaccine hesitancy remains high. [18]

An observational cohort study reported that two doses of the BNT162b2 mRNA vaccine was 89% effective for preventing COVID-19 related hospital admissions in pregnant women during the wild type and Alpha variant dominant periods in Israel, which was similar to the estimated efficacy in the general population. [19] In another study from Israel, two or three mRNA vaccine doses were 96% and 99% effective, respectively, in preventing severe disease in pregnant women during the Delta period, decreasing to 83% and 94% during the Omicron period. [20] However, large-scale studies directly comparing vaccine effectiveness between pregnant and non-pregnant women of reproductive age at the population level after adjusting for sociodemographic characteristics linked with severe illness and vaccine uptake are lacking.

In this study, we used population-level linked administrative data for England to estimate vaccine effectiveness for preventing COVID-19 hospital admission among women who were infected with SARS-CoV-2 during pregnancy, compared to women who were not pregnant when they were infected.

## Methods

### Study data

We conducted a population-based cohort study using data from the Office for National Statistics (ONS) Public Health Data Asset (PHDA). The ONS PHDA is a linked dataset combining: the 2011 Census; mortality records; the General Practice Extraction Service (GPES) Data for Pandemic Planning and Research (GDPPR); Hospital Episode Statistics (HES); vaccination data from the National Immunisation Management System (NIMS); and National Health Service (NHS) Test and Trace Pillar 1 (swab testing for the virus in UK Health Security Agency labs and NHS hospitals for those with a clinical need, and health and care workers) and Pillar 2 (swab testing for the virus in the wider population, through commercial partnerships, either processed in a lab or more rapidly via lateral flow device tests) data. [21]

To obtain NHS numbers, the 2011 Census was linked to the 2011 to 2013 NHS Patient Registers using deterministic and probabilistic matching, with an overall linkage rate of 94.6% (detailed description of the linkage methodology and quality evaluation have been previously reported [22]). Further linkage to deaths registrations data, GDPPR, HES, and NIMS data was performed deterministically using a unique identifier (NHS number).

We linked the PHDA to NHS birth notifications data for 2020, 2021, and January to March of 2022 using mothers’ NHS numbers. The birth notification is a document completed by the doctor or midwife present at the birth. It is used to notify registration offices of the birth and issue NHS numbers to babies. Birth notifications data only include pregnancies resulting in a live birth or stillbirth after at least 24 weeks of gestation. There are small differences in the number of births recorded between birth notifications and registrations data, but the two data sources are very similar. [23]

We used data from the 2021 Census to derive more up-to-date sociodemographic characteristics for participants in the study. The 2021 Census was deterministically linked to the NHS Personal Demographics Service to retrieve NHS numbers, with a linkage rate of 94.6%. Following clerical review of links made, the precision (proportion of true links) was estimated to be 99.4% (95% confidence interval 96.5% to 100.0%); 1.6% of these links involved multiple 2021 Census records linked to the same NHS number, which were excluded following deduplication. The 2021 Census was then linked to the PHDA using NHS numbers.

### Study population and design

The study cohort comprised women who had a first recorded SARS-CoV-2 infection between 8^th^ December 2020 (the start of the vaccination campaign in the UK) and 31^st^ August 2021 (with no evidence of prior infection) and: (i) enumerated at the 2011 Census and living in a private household; (ii) aged 18 to 45 years at the start of the study period; (iii) could be linked to the 2011 to 2013 NHS Patient Registers; (iv) could be linked to at least one GDPPR record; and (v) resident in England according to most recent postcodes held in GDPPR.

The index date for the start of follow-up was the earliest evidence of SARS-CoV-2 infection within the study period. Evidence of SARS-CoV-2 infection was determined by a positive swab for SARS-CoV-2 using a polymerase chain reaction (PCR) test or lateral flow device (LFD) recorded in NHS Test and Trace data, or a hospital inpatient admission or outpatient appointment with an ICD-10 code for U07.1 (COVID-19, virus identified) or U07.2 (COVID-19, virus not identified) as the primary or secondary diagnosis.

### Method for identifying pregnancies

Two data sources were used to identify pregnancy status at time of SARS-CoV-2 infection. NHS birth notifications data were used to identify women who were pregnant when infected with SARS-CoV-2 and went on to have a live birth or stillbirth. HES data were used to identify women who were pregnant when they were infected with SARS-CoV-2 but for whom a birth notification was not recorded (e.g., pregnancies that ended before 24 weeks, which are not recorded in birth notifications data, or pregnancies that were ongoing at the end of the study period). See **Figure 1** for an overview of the methodology; a more detailed description is available in the Supplementary Methods.

**Figure 1.**
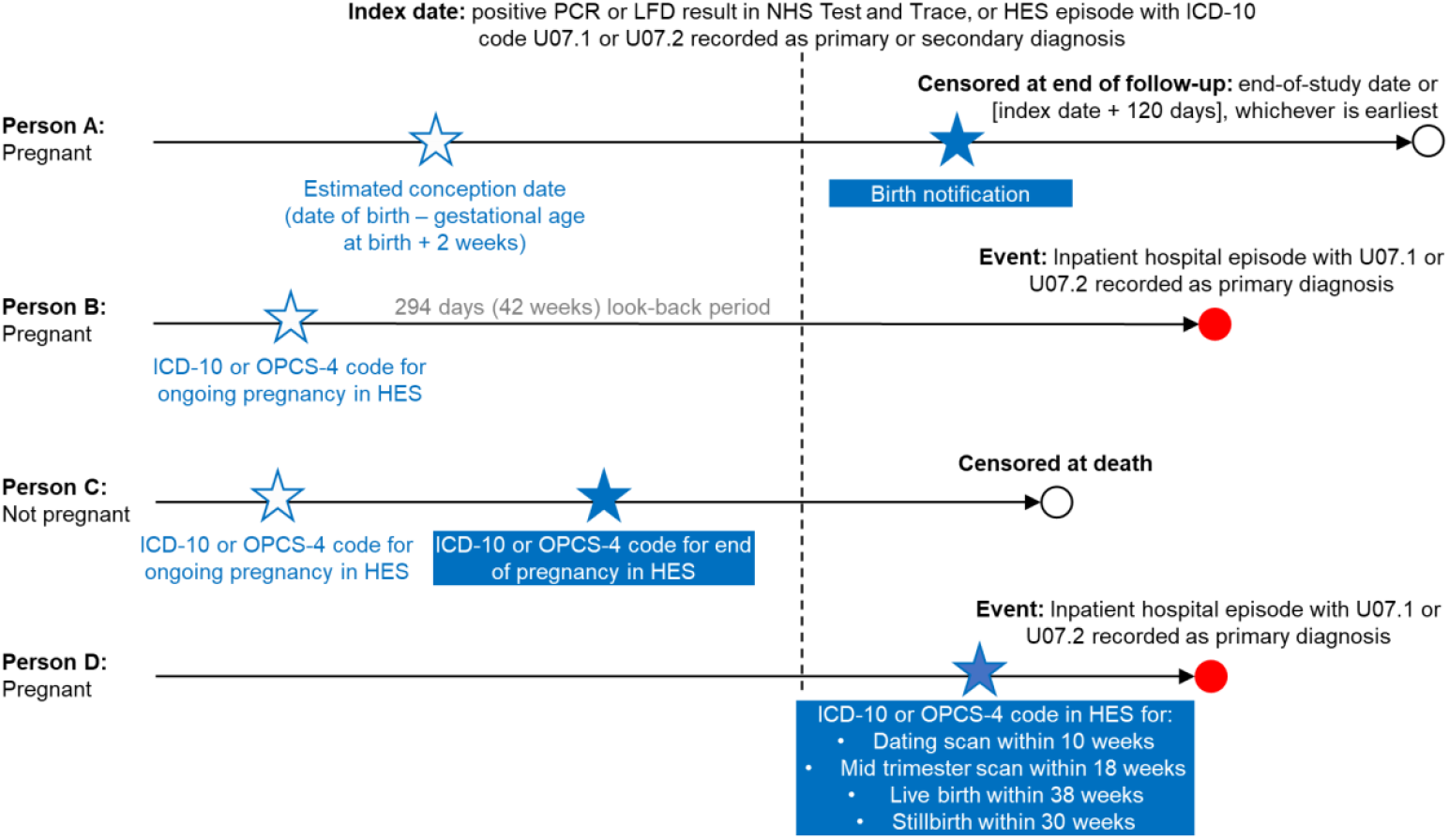
Method for identifying women who were pregnant when infected with SARS-CoV-2.

### Exposure

The exposure was vaccination status derived from NIMS data. Vaccination status was defined as the number of doses received at least 14 days prior to the index date. Participants were classified as single-vaccinated if they had received one dose of a COVID-19 vaccine at least 14 days before infection, or double-vaccinated if they had received two doses of a COVID-19 vaccine at least 14 days before infection.

### Covariates

We adjusted for sociodemographic factors known from previous studies to be associated with risk of severe COVID-19 outcomes and vaccine uptake. [24, 25, 26, 27, 28] For 91.6% of participants, the following covariates were included from the 2021 Census: age, ethnic group, English language proficiency, country of birth, keyworker status, highest qualification held, disability status, and health status (**Table S1**). For the remaining 8.4% of participants that could not be linked to the 2021 Census, these variables were based on 2011 Census data. Missing Census responses were imputed using nearest-neighbour donor imputation. [29]

Geographical covariates were derived from postcodes in GDPPR data. Region and Rural/Urban classification were retrieved from the National Statistics Postcode Lookup. [30] Index of Multiple Deprivation (IMD) was retrieved from the English Indices of Deprivation, 2019. [31]

### Outcome

The outcome was a hospital inpatient episode with an ICD-10 code for U07.1 or U07.2 recorded as the primary diagnosis and occurring within 120 days of the index date; this time frame was used to avoid inclusion of outcomes related to a subsequent infection episode. [32]

### Statistical analysis

We calculated age-standardised rates of COVID-19 hospital admission (per 100,000 infections) by vaccination status and pregnancy status, standardised to the 2013 European Standard Population. [33]

We used Cox proportional hazards models to assess how the rate of COVID-19 hospital admission varied by vaccination status (reference group: unvaccinated). Models were stratified by pregnancy status at the time of first SARS-CoV-2 infection. Follow-up time was calculated from first infection (index date) until COVID-19 hospital admission, death, or 120 days of follow-up, whichever occurred first. The proportional hazards assumption was assessed by inspecting plots of the Schoenfeld residuals. We calculated hazard ratios adjusted for all sociodemographic covariates and calendar time of infection to account for differences in COVID-19 variant, changes in treatment strategies, and changes in hospital capacity over the study period (**Table S1** describes how variables were modelled). Vaccine effectiveness (VE) was calculated as the complement of the hazard ratio.

To assess differential waning of VE between single- and double-vaccinated women, we stratified the analysis by time since last vaccine dose (14 to 90 days versus more than 90 days [33, 34]).

We conducted sensitivity analysis excluding women who were infected with SARS-CoV-2 before 11^th^ June 2021 (42 weeks before 31^st^ March 2022, the most recent birth notifications data available) who were identified as pregnant in HES data, but not in birth notifications data. These women may have been pregnant previously but were no longer when they were infected due to early end of pregnancy that was not recorded in HES records or birth notifications data.

All statistical analyses were conducted using R version 3.5. Cox proportional hazards models were implemented using the survival package (version 2.41-3). [36]

### Patient and public involvement

We did not directly involve patients and the public in the design and conception of the study, primarily because of the pace at which this study was conducted to inform the UK government’s response to the COVID-19 pandemic.

## Results

### Characteristics of the study population

The study population included 815,477 women aged 18 to 45 years (mean age, 30.4 years; standard deviation (SD), 8.1 years); 33,549 (4.1%) were identified as pregnant when they were infected with SARS-CoV-2 (**Table 1**).

**Table 1.**
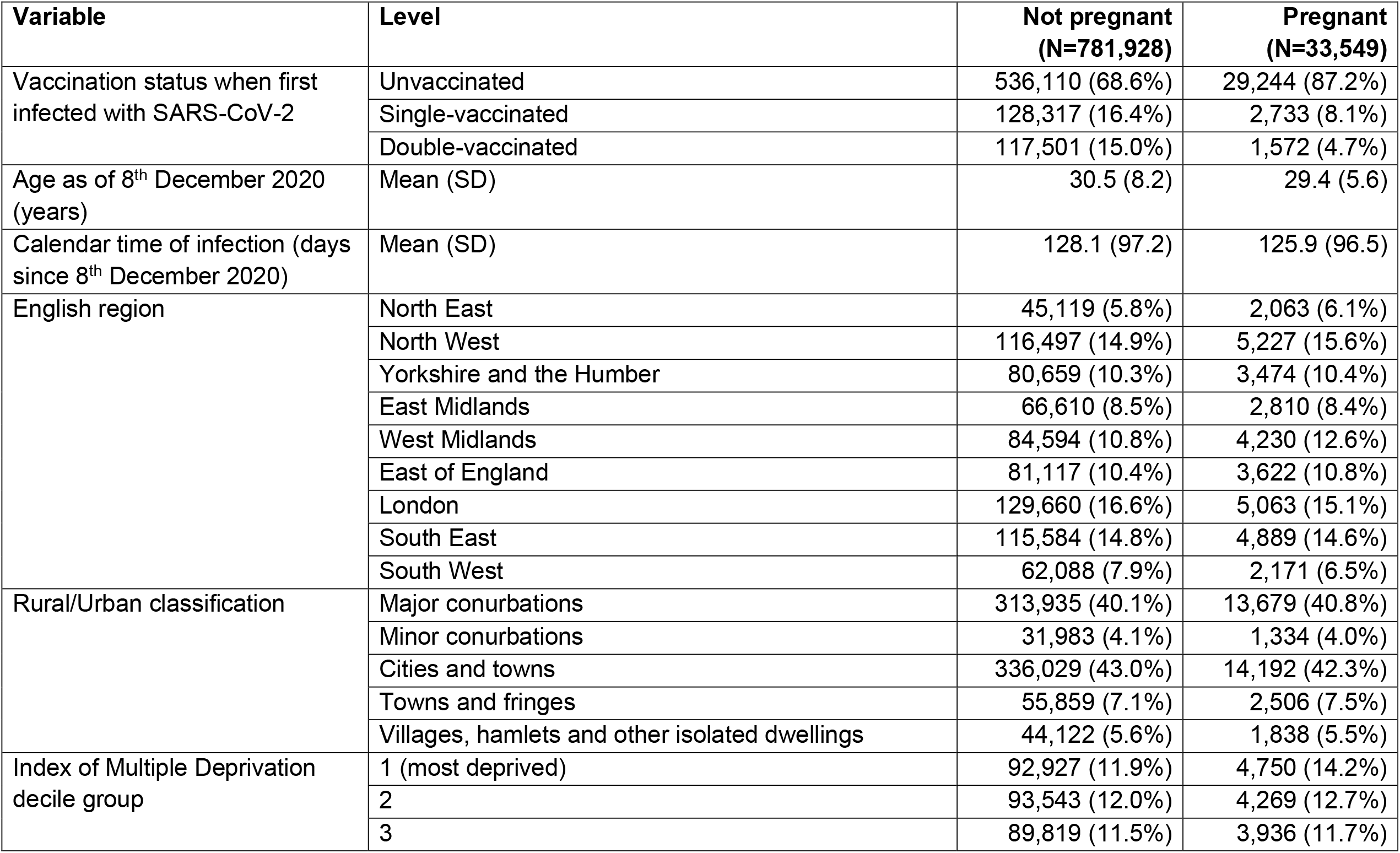
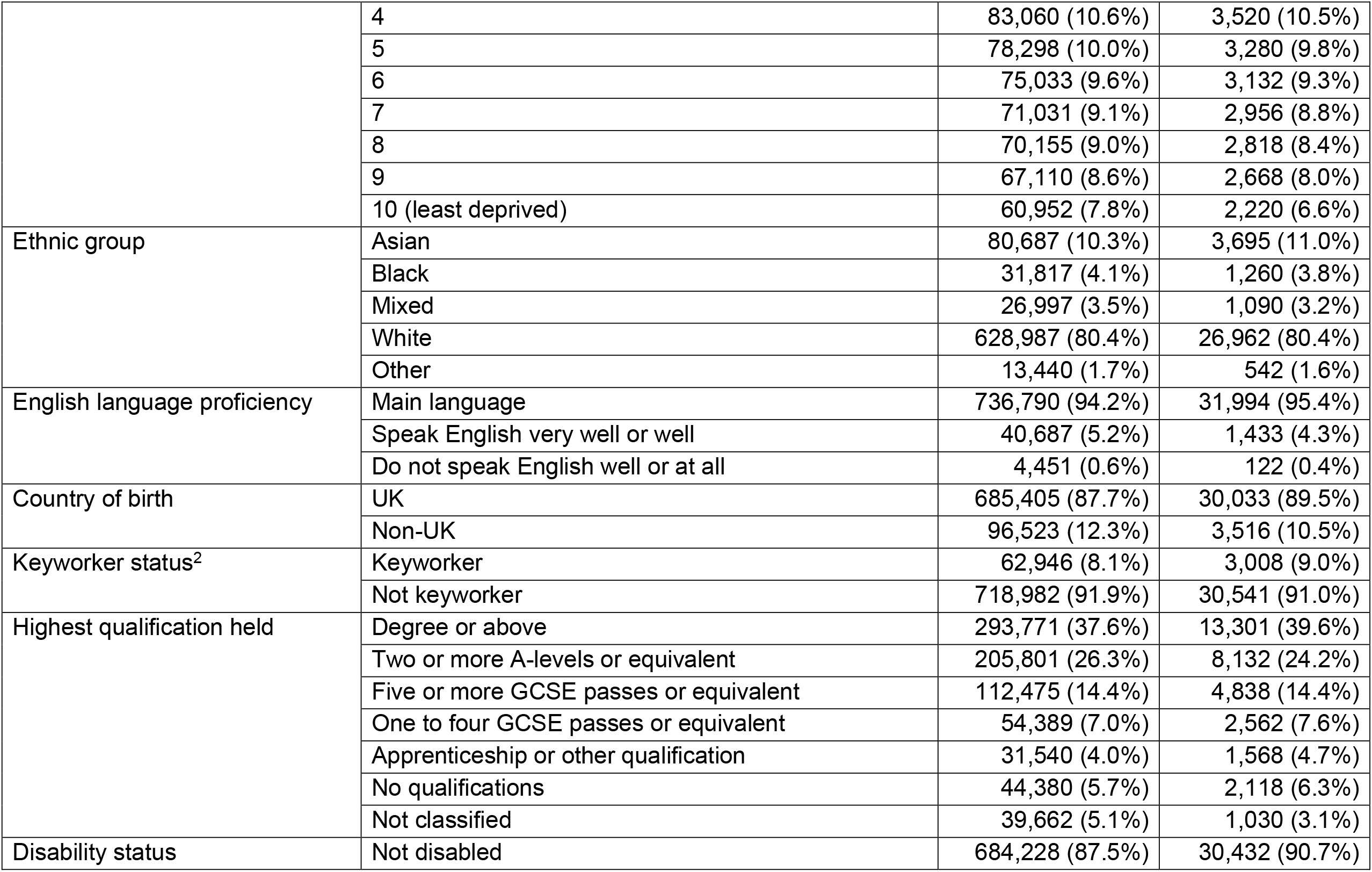
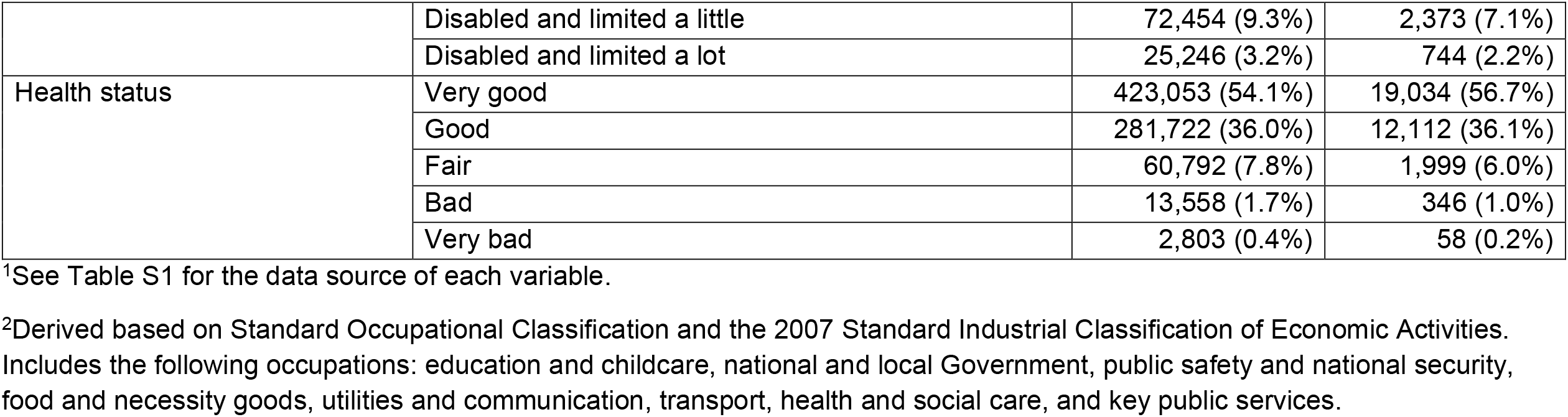
Demographic characteristics of the study population^1^

Among women identified as pregnant at the time of SARS-CoV-2 infection, 87.2% were unvaccinated, 8.1% were single-vaccinated, and 4.7% were double-vaccinated. Among non-pregnant women, 68.6% were unvaccinated, 16.4% were single-vaccinated, and 15.0% were double-vaccinated.

### Age-standardised rates of COVID-19 hospital admission

Overall, 9,889 COVID-19 hospital admissions occurred in the study period: 1,895 (19.2%) were among women who were pregnant when infected (**Table 2**), of whom 1,807 (95.4%) were unvaccinated; and 7,994 (80.8%) occurred in non-pregnant women, of whom 7,028 (87.9%) were unvaccinated.

**Table 2.**
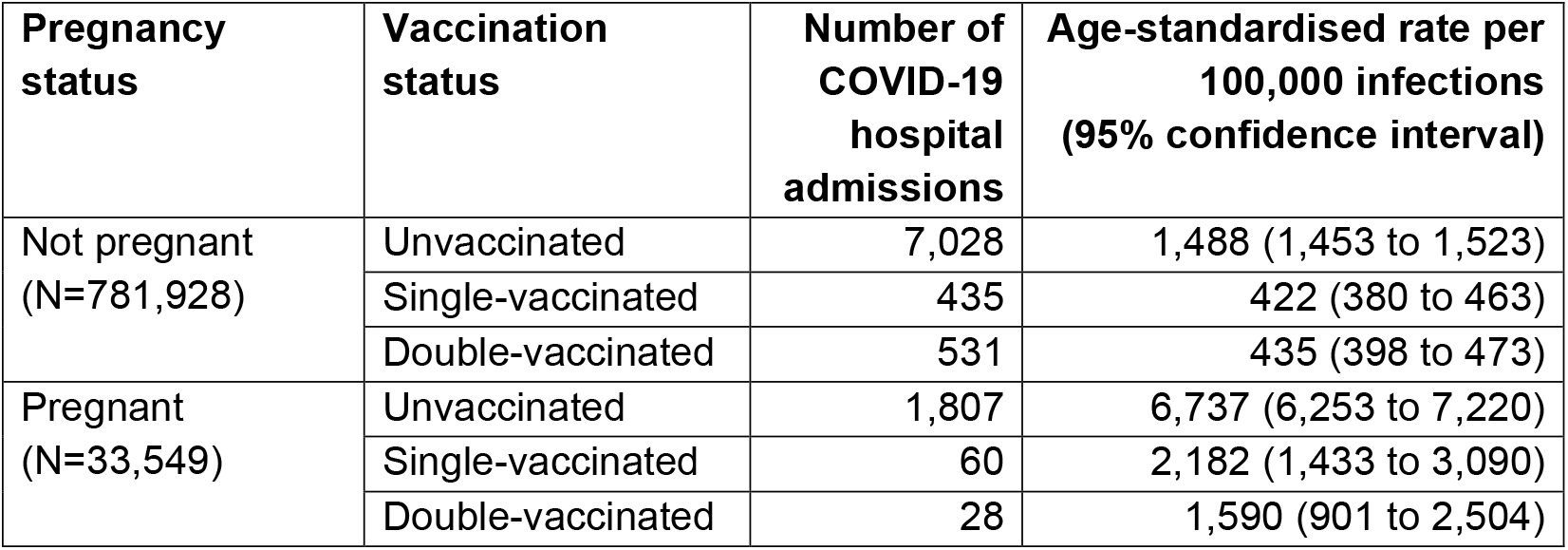
Number and age-standardised rates (per 100,000 infections) of COVID-19 hospital admissions by pregnancy status and vaccination status when first infected with SARS-CoV-2.

For both pregnant and non-pregnant women, age-standardised rates of COVID-19 hospital admission (per 100,000 infections) were higher among those who were unvaccinated compared with those who were single- or double-vaccinated (**Table 2**). Among pregnant women, the age-standardised rate of COVID-19 hospital admission was 6,737 (95% confidence interval (CI): 6,253 to 7,220) per 100,000 infections for those who were unvaccinated, 2,182 (1,433 to 3,090) for those who were single-vaccinated, and 1,560 (901 to 2,504) for those who were double-vaccinated. The corresponding rates for non-pregnant women were 1,488 (1,453 to 1,523), 422 (380 to 463), and 435 (398 to 473), respectively.

### Vaccine effectiveness for preventing COVID-19 hospital admission

Compared with unvaccinated pregnant women, VE for preventing COVID-19 hospital admission was 76% (95% CI: 69% to 82%) for single-vaccinated pregnant women and 83% (75% to 88%) for double-vaccinated pregnant women (**Figure 2**). Corresponding estimates for non-pregnant women were 79% (76% to 81%) and 82% (80% to 83%), respectively.

**Figure 2.**
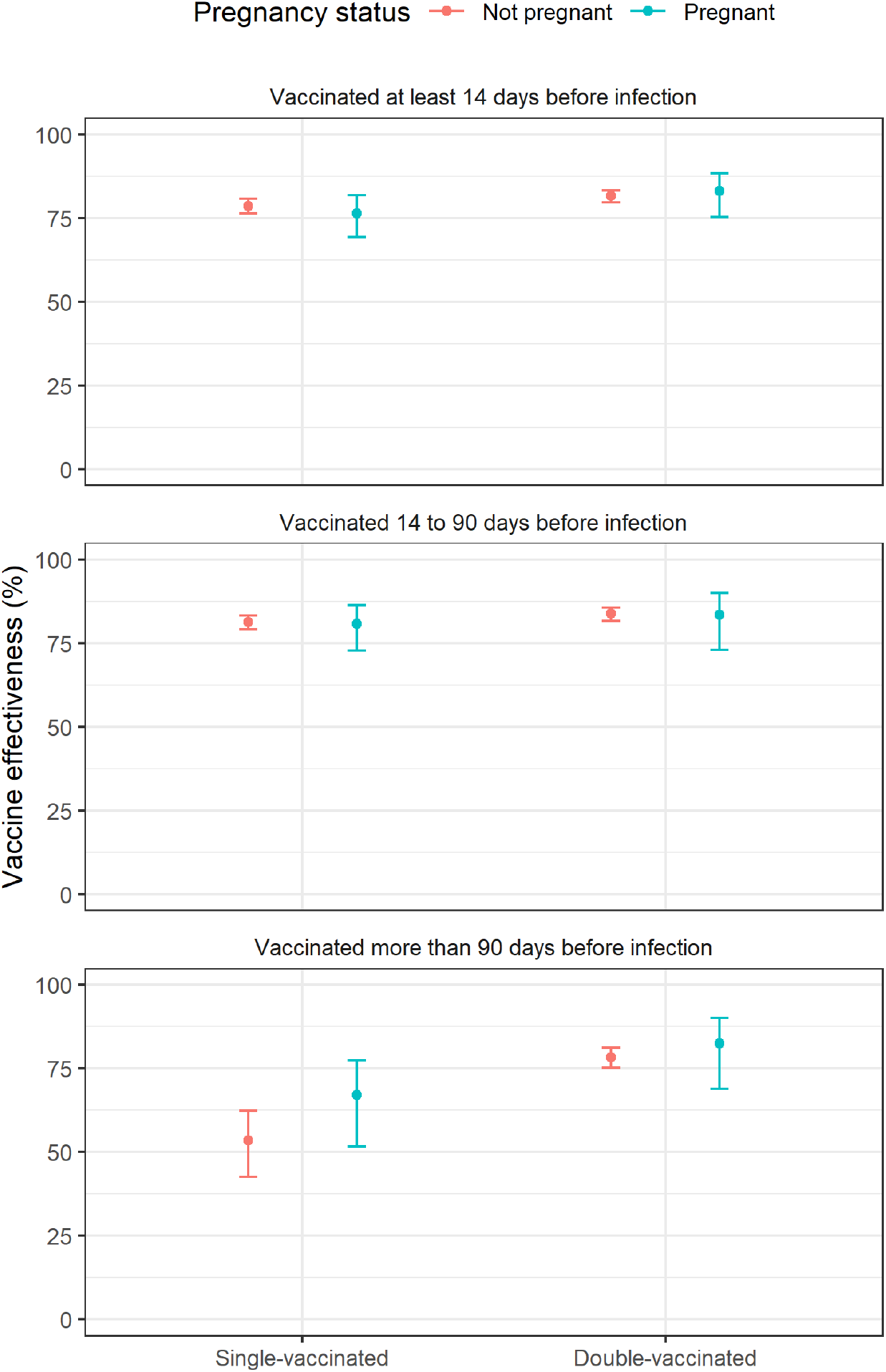
Vaccine effectiveness for preventing COVID-19 hospital admission, stratified by pregnancy status and time since vaccination. Estimates were calculated as the complement of the hazard ratio from Cox proportional hazards models adjusted for age, calendar time of infection, region, Index of Multiple deprivation decile group, Rural/Urban classification, ethnic group, English language proficiency, country of birth, keyworker status, highest qualification held, disability status, and health status.

Inspection of Schoenfeld residuals plots indicated potential non-proportional hazards for vaccination after approximately 4 weeks of follow-up (**Figure S1**). We therefore conducted a sensitivity analysis restricted to 28 days of follow-up and the results were not substantially different (**Table S2**). Results were also robust to excluding 1,671 women who were potentially misclassified as pregnant from the analysis (**Table S3**).

In both pregnant and non-pregnant women VE was similar between those single-vaccinated or double-vaccinated 14 to 90 days before first infection (**Figure 2**). Among those vaccinated more than 90 days before first infection, VE was lower for single-vaccinated non-pregnant women (54% [43% to 62%]) than double-vaccinated non-pregnant women (78% [75% to 81%]) (*p*<0.001). A similar pattern was observed in pregnant women (single-vaccinated 67% [52% to 77%]; double-vaccinated 82% [69% to 90%]) (*p*=0.07).

## Discussion

### Summary of main findings

Our study shows that vaccination against COVID-19 was associated with a reduced risk of COVID-19 hospital admission during the Alpha and Delta periods in women of reproductive age, regardless of their pregnancy status when infected with SARS-CoV-2. The effectiveness of COVID-19 vaccines against hospitalisation waned more quickly after one dose than two doses in both pregnant and non-pregnant women.

### Comparison with other studies

Our findings are consistent with other studies showing that COVID-19 vaccination is effective in reducing the risk of severe illness in pregnant women infected with SARS-CoV-2. [13, 14] Dagan et al. (2021) found that two doses of the BNT162b2 mRNA vaccine was 89% effective for preventing COVID-19 related hospitalisation in pregnant women during the wild type and Alpha dominant periods in Israel. [19] Another study from Israel found that two doses of the BNT162b2 mRNA vaccine were 61% effective at preventing hospitalisation with confirmed SARS-CoV-2 infection and 96% effective at preventing severe disease among pregnant women during the Delta period. [20] We found that two vaccine doses were 83% effective for preventing hospital admissions where COVID-19 was recorded as the primary condition being treated (or was responsible for the primary condition being treated) in women who were pregnant when infected with SARS-CoV-2 during the Alpha and Delta dominant periods combined. This was after adjusting for a range of sociodemographic characteristics associated with risk of severe illness and vaccine uptake.

We also found that having two doses of a COVID-19 vaccine was associated with greater protection against COVID-19 hospital admission than one dose among those vaccinated more than 90 days before infection, suggesting faster waning of vaccine effectiveness following a single vaccine dose. Our study did not assess effectiveness against the Omicron variant or the effectiveness of booster vaccines. However, previous evidence suggests that three vaccine doses are more effective than two doses for preventing severe illness in pregnant women infected during the Omicron period. [20] None of the pregnant women admitted to intensive care for COVID-19 in the UK during the Omicron dominant variant period had received three vaccine doses. [37] Considering evidence in the general population, that the effectiveness of three doses for preventing severe illness following infection with Omicron wanes over time, [37, 38] future research should compare the waning of third dose effectiveness between pregnant and non-pregnant women.

### Strengths and limitations

The main strength of our study derives from using a nationwide linked data asset combining the 2011 and 2021 Censuses, mortality records, hospital records, birth notifications data, vaccinations data, and SARS-CoV-2 testing data from national testing programmes. Using hospital data and birth notifications data, we were able to identify women that were pregnant when they were infected with SARS-CoV-2. We adjusted for a range of sociodemographic characteristics associated with vaccine uptake and risk of severe COVID-19 outcomes. For most participants, these variables were based on up-to-date data from the 2021 Census.

A limitation of our study is that the study population may not fully represent the at-risk population. The cohort does not include people living in England in 2011 who did not participate in the 2011 Census (estimated to be 5% of households [39]); those who could not be linked to the 2011 to 2013 NHS Patient Registers; those who immigrated since 2011; or those not registered with a general practitioner at the start of the coronavirus pandemic.

Misclassification of pregnancy status is possible because of limited data availability, especially in the first trimester and for women whose pregnancy ended before 24 weeks. Consequently, women who were pregnant when infected with SARS-CoV-2 who had short pregnancies may have been misclassified as not pregnant.

Conversely, women who had been pregnant but were no longer so when they were infected with SARS-CoV-2 may have been incorrectly classified as being pregnant. This could bias results towards showing the pregnant and non-pregnant groups to be more similar than they actually are in terms vaccine effectiveness. However, this is likely to have introduced very limited bias as we found similar results in a sensitivity analysis excluding participants who were potentially misclassified as pregnant.

People with asymptomatic COVID-19 may be less likely to seek a test for SARS-CoV-2 via the NHS Test and Trace programme. They may also be less likely to report the result of the test. Consequently, COVID-19 hospital admission rates in the study population may overestimate the true rates in the general population as asymptomatic infections are likely to be under-represented. More asymptomatic infections may have been detected in the pregnant group due to a combination of altered test-seeking behaviour in pregnancy and the requirement to undergo testing prior to antenatal appointments.

COVID-19 hospital admissions were defined as inpatient admissions, where COVID-19 was recorded as the primary diagnosis. However, this will include some hospital admissions where the initial reason for admission was not related to COVID-19, but the patient was subsequently diagnosed with, and treated for, COVID-19 while in hospital. This will include SARS-CoV-2 infections that occur before admission to hospital and hospital acquired infections, which are probably more common among pregnant women who are more likely to have hospital contact than women in the general population.

### Implications

Pregnant women were identified as a vulnerable group and prioritised for COVID-19 vaccination in December 2021 by the Joint Committee on Vaccination and Immunisation (JCVI) in the UK. The Royal College of Obstetricians and Gynaecologists strongly recommend that COVID-19 vaccines are offered to all pregnant women. [41] Other studies have shown a lower risk of stillbirth in those vaccinated, and no evidence for adverse pregnancy outcomes following COVID-19 vaccination. [12, 42] Vaccination coverage among women giving birth has been increasing over time, but uptake remains lower at the time of delivery in women from ethnic minority groups, with lowest vaccination rates in black women, and those living in deprived areas. [11] Interventions to address these inequalities and engagement with pregnant women to ensure uptake of future booster vaccines are needed, since many women who become pregnant may have received their last dose of a vaccine several months previously.

### Conclusions

During the Alpha and Delta periods, COVID-19 vaccination was associated with a reduced risk of COVID-19 hospital admission in women infected with SARS-CoV-2 during pregnancy as well as among non-pregnant women. Increasing vaccine uptake in pregnant women may contribute to reduced levels of avoidable harms to pregnant women associated with COVID-19. These data add to the evidence base regarding the protective effect of vaccination for women infected with SARS-CoV-2 during pregnancy by providing real-world evidence in a population which was originally excluded from the vaccine trials.

## Data Availability

In accordance with NHS Digital's Information Governance requirements, the study data cannot be shared.

## Acknowledgments

We would like to thank Dr Clara Calvert (University of Edinburgh) for her helpful comments on the manuscript.

## Contributors

MLB, CH, DA, and VN conceptualised and designed the study. MLB and RS prepared the study data. MLB performed the statistical analysis. LC and CH quality assured the underlying data and results. All authors contributed to interpretation of the findings. MLB and CH wrote the original draft. All authors contributed to review and editing of the manuscript and approved the final version.

## Ethical approval

Ethical approval was obtained from the National Statistician’s Data Ethics Advisory Committee (NSDEC(20)12).

## Transparency declaration

The lead author affirms that the manuscript is an honest, accurate, and transparent account of the study being reported; that no important aspects of the study have been omitted; and that any discrepancies from the study as planned (and, if relevant, registered) have been explained.

## Dissemination declaration

The use of deidentified data precludes direct dissemination to participants.

## Supplementary Materials

### Method for identifying pregnancy from birth notifications data

The estimated conception date was calculated by subtracting gestational age from the date of birth on the earliest birth notification occurring after the index date. Two weeks were added to account for the fact that gestational age is defined from the start of the last menstrual period, and conception is assumed to occur at the mid-way point (day 14) of a typically assumed average 28-day menstrual cycle. Women were classified as pregnant at the index date if there was evidence of SARS-CoV-2 infection between the estimated conception date and date of birth on the birth notification.

Gestational age was missing for 0.3% of the birth notifications used to estimate the conception date. For these records, gestational age was imputed as 40 weeks for live births and 32 weeks for stillbirths (the approximate average length of gestation for these outcomes, calculated from published births data [43]).

### Method for identifying pregnancy from Hospital Episode Statistics (HES) data

A. We searched HES data for hospital episodes with evidence of ongoing pregnancy and end of pregnancy in the 42 weeks before the index date (**Tables S4** to **S7** report the code lists used). Participants were classified as pregnant at the index date if they met all of the following criteria:
  - They had at least one hospital episode with an ICD-10 or Classification of Interventions and Procedures version 4 (OPCS-4) code indicating pregnancy over the 42 weeks prior to and including the index date.
  - The most recent ICD-10 or OPCS-4 code over the 42 weeks prior to the index date indicated ongoing pregnancy (not the end of pregnancy).
  - There was no end of pregnancy code in the six weeks prior to the most recent ongoing pregnancy code (some of the ongoing pregnancy codes relate to conditions that can be diagnosed in the post-partum period, up to six weeks after delivery).
B. We also searched HES data for hospital episodes with evidence of ongoing pregnancy (routine obstetric scans only) and birth events occurring after the index date. Participants were classified as pregnant at the index date if any of the following criteria were met:
  - They had a hospital episode with an OPCS-4 code for a dating scan (R36.1; normally performed at week 12 of pregnancy) up to 70 days after the index date.
  - They had a hospital episode with an OPCS-4 code for a mid-trimester scan (R36.3; normally performed at week 20 of pregnancy) up to 126 days after the index date.
  - They had a hospital episode with an ICD-10 code for a live birth (Z37.0, Z37.2, Z37.5 or Z38) or mixed live and stillbirth (Z37.3 or Z37.6) up to 38 weeks after the index date.
  - They had a hospital episode with an ICD-10 code for a stillbirth (Z37.1, Z37.4 or Z37.7) up to 30 weeks after the index date.

**Table S1.**
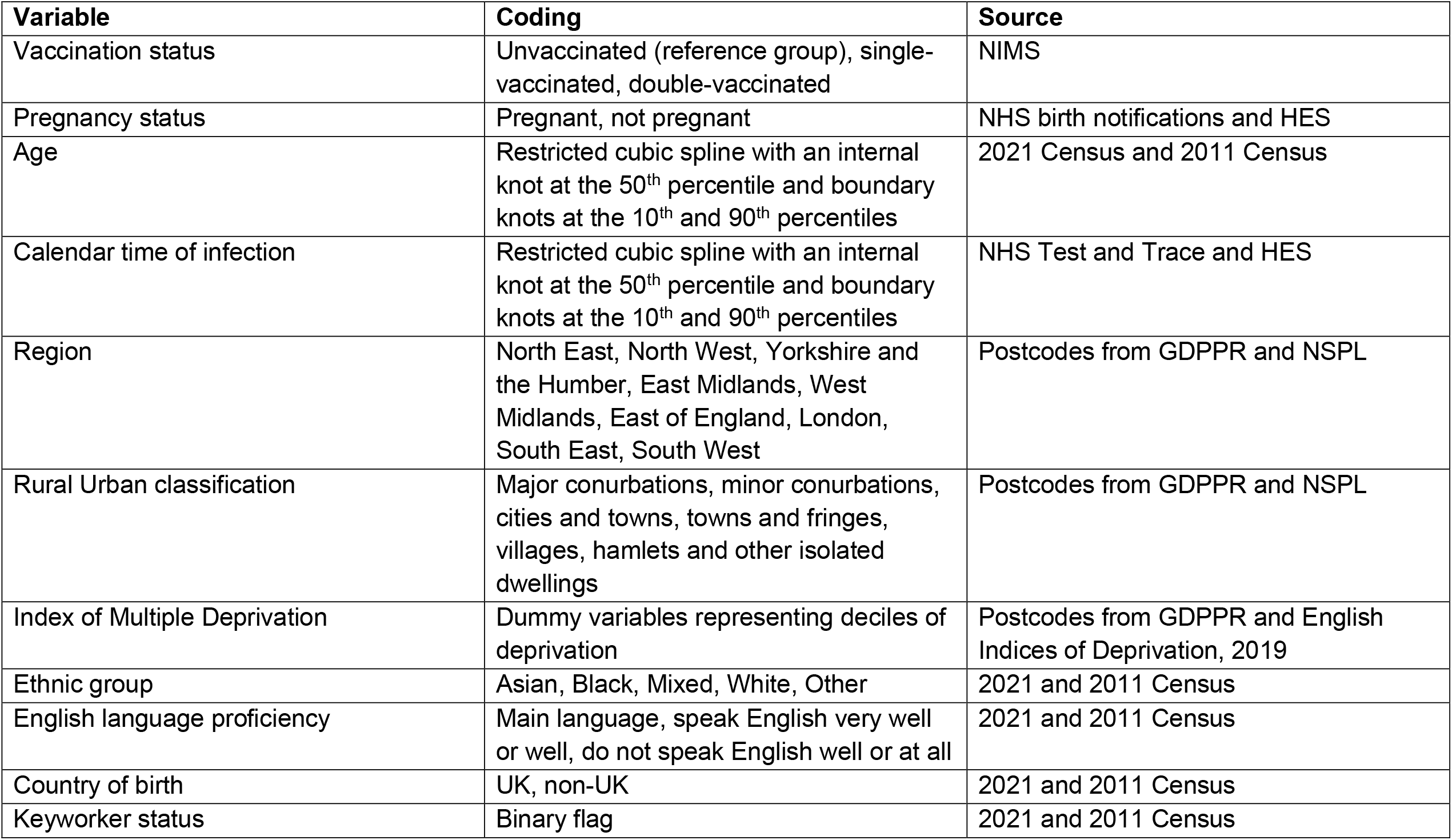
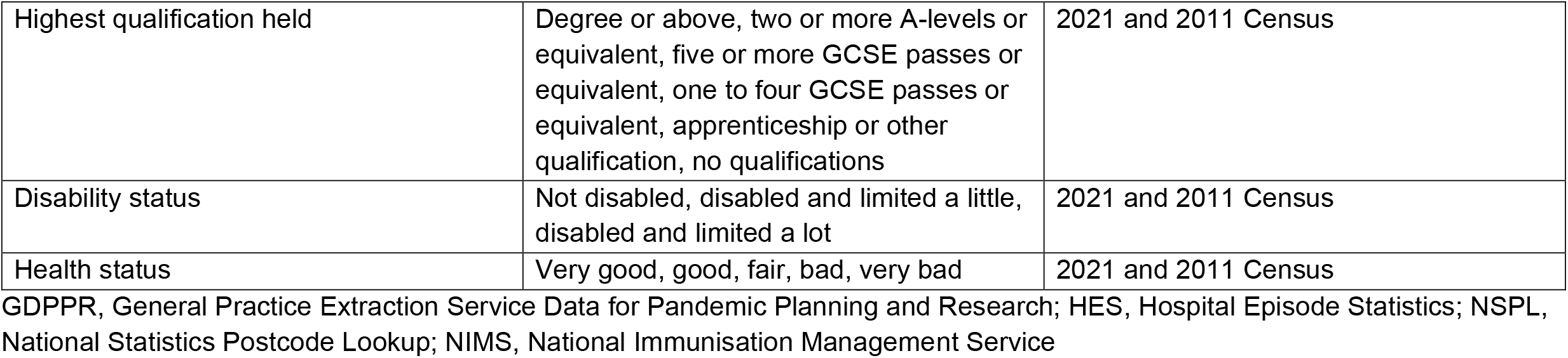
Coding and source of variables included in the analysis

| **Variable** | **Coding** | **Source** |
| --- | --- | --- |
| Vaccination status | Unvaccinated (reference group), single-vaccinated, double-vaccinated | NIMS |
| Pregnancy status | Pregnant, not pregnant | NHS birth notifications and HES |
| Age | Restricted cubic spline with an internal knot at the 50<sup>th</sup> percentile and boundary knots at the 10<sup>th</sup> and 90<sup>th</sup> percentiles | 2021 Census and 2011 Census |
| Calendar time of infection | Restricted cubic spline with an internal knot at the 50<sup>th</sup> percentile and boundary knots at the 10<sup>th</sup> and 90<sup>th</sup> percentiles | NHS Test and Trace and HES |
| Region | North East, North West, Yorkshire and the Humber, East Midlands, West Midlands, East of England, London, South East, South West | Postcodes from GDPPR and NSPL |
| Rural Urban classification | Major conurbations, minor conurbations, cities and towns, towns and fringes, villages, hamlets and other isolated dwellings | Postcodes from GDPPR and NSPL |
| Index of Multiple Deprivation | Dummy variables representing deciles of deprivation | Postcodes from GDPPR and English Indices of Deprivation, 2019 |
| Ethnic group | Asian, Black, Mixed, White, Other | 2021 and 2011 Census |
| English language proficiency | Main language, speak English very well or well, do not speak English well or at all | 2021 and 2011 Census |
| Country of birth | UK, non-UK | 2021 and 2011 Census |
| Keyworker status | Binary flag | 2021 and 2011 Census |
| Highest qualification held | Degree or above, two or more A-levels or equivalent, five or more GCSE passes or equivalent, one to four GCSE passes or equivalent, apprenticeship or other qualification, no qualifications | 2021 and 2011 Census |
| Disability status | Not disabled, disabled and limited a little, disabled and limited a lot | 2021 and 2011 Census |
| Health status | Very good, good, fair, bad, very bad | 2021 and 2011 Census |

GDPPR, General Practice Extraction Service Data for Pandemic Planning and Research; HES, Hospital Episode Statistics; NSPL, National Statistics Postcode Lookup; NIMS, National Immunisation Management Service

**Table S2.** Hazard ratios for COVID-19 hospital admission within 28 days of SARS-CoV-2 infection

| **Pregnancy status** | **Vaccination status** | **Hazard ratio<sup>1</sup> (95% CI)** |
| --- | --- | --- |
| Not pregnant | Single-vaccinated | 0.21 (0.19 to 0.24) |
|  | Double-vaccinated | 0.18 (0.16 to 0.20) |
| Pregnant | Single-vaccinated | 0.24 (0.18 to 0.31) |
|  | Double-vaccinated | 0.18 (0.12 to 0.26) |

<sup>1</sup>Results obtained from Cox proportional hazards models adjusted for age, calendar time of infection, region, Index of Multiple deprivation decile group, Rural/Urban classification, ethnic group, English language proficiency, country of birth, keyworker status, highest qualification held, disability status, and health status.

**Table S3.** Hazard ratios for COVID-19 hospital admission in pregnant women from sensitivity analysis excluding those who were potentially misclassified as pregnant

| **Vaccination status** | **Hazard ratio<sup>1</sup> (95% CI)** |
| --- | --- |
| Single-vaccinated | 0.23 (0.18 to 0.31) |
| Double-vaccinated | 0.16 (0.11 to 0.24) |

<sup>1</sup>Results obtained from Cox proportional hazards models adjusted for age, calendar time of infection, region, Index of Multiple deprivation decile group, Rural/Urban classification, ethnic group, English language proficiency, country of birth, keyworker status, highest qualification held, disability status, and health status.

**Table S4.** ICD-10 code list for ongoing pregnancy

| Code | Description |
| --- | --- |
| O07% | Failed attempted abortion |
| O10% | Pre-existing hypertension complicating pregnancy, childbirth and the puerperium |
| O11% | Pre-eclampsia superimposed on chronic hypertension |
| O12% | Gestational oedema and proteinuria without hypertension |
| O13% | Gestational hypertension |
| O14% | Pre-eclampsia |
| O15% | Eclampsia |
| O16% | Unspecified maternal hypertension |
| O20% | Haemorrhage in early pregnancy |
| O21% | Excessive vomiting in pregnancy |
| O22% | Venous complications and haemorrhoids in pregnancy |
| O23% | Infections of genitourinary tract in pregnancy |
| O24% | Diabetes mellitus in pregnancy |
| O25% | Malnutrition in pregnancy |
| O26% | Maternal care for other conditions predominantly related to pregnancy |
| O28% | Abnormal findings on antenatal screening of mother |
| O29% | Complications of anaesthesia during pregnancy |
| O30% | Multiple gestation |
| O31% | Complications specific to multiple gestation |
| O32% | Maternal care for known or suspected malpresentation of fetus |
| O33% | Maternal care for known or suspected disproportion |
| O34% | Maternal care for known or suspected abnormality of pelvic organs |
| O35% | Maternal care for known or suspected fetal abnormality and damage |
| O36% | Maternal care for other known or suspected fetal problems |
| O40% | Polyhydramnios |
| O41% | Other disorders of amniotic fluid and membranes |
| O42% | Premature rupture of membranes |
| O43% | Placental disorders |
| O44% | Placenta praevia |
| O45% | Premature separation of placenta [abruptio placentae] |
| O46% | Antepartum haemorrhage, not elsewhere classified |
| O47% | False labour |
| O48% | Prolonged pregnancy |
| O88% | Obstetric embolism |
| O98% | Maternal infectious and parasitic diseases classifiable elsewhere but complicating pregnancy, childbirth and the puerperium |
| O99% | Other maternal diseases classifiable elsewhere but complicating pregnancy, childbirth and the puerperium |
| Z321 | Pregnancy confirmed |
| Z33% | Pregnant state, incidental |
| Z34% | Supervision of normal pregnancy |
| Z35% | Supervision of high-risk pregnancy |
| Z36% | Antenatal screening |

**Table S5.**
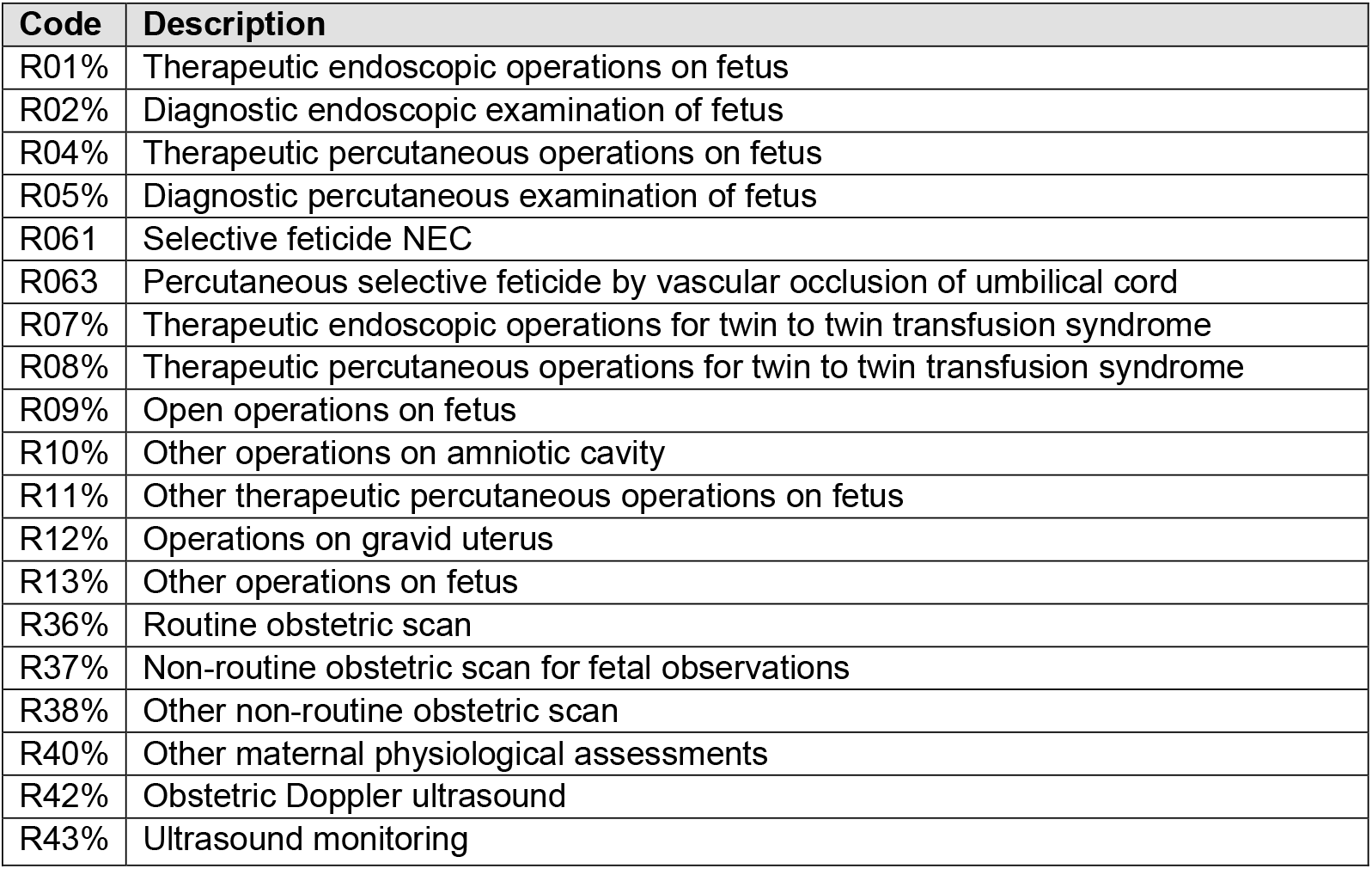
OPCS-4 code list for ongoing pregnancy

| Code | Description |
| --- | --- |
| R01% | Therapeutic endoscopic operations on fetus |
| R02% | Diagnostic endoscopic examination of fetus |
| R04% | Therapeutic percutaneous operations on fetus |
| R05% | Diagnostic percutaneous examination of fetus |
| R061 | Selective feticide NEC |
| R063 | Percutaneous selective feticide by vascular occlusion of umbilical cord |
| R07% | Therapeutic endoscopic operations for twin to twin transfusion syndrome |
| R08% | Therapeutic percutaneous operations for twin to twin transfusion syndrome |
| R09% | Open operations on fetus |
| R10% | Other operations on amniotic cavity |
| R11% | Other therapeutic percutaneous operations on fetus |
| R12% | Operations on gravid uterus |
| R13% | Other operations on fetus |
| R36% | Routine obstetric scan |
| R37% | Non-routine obstetric scan for fetal observations |
| R38% | Other non-routine obstetric scan |
| R40% | Other maternal physiological assessments |
| R42% | Obstetric Doppler ultrasound |
| R43% | Ultrasound monitoring |

**Table S6.** ICD-10 code list for the end of pregnancy outcomes

| **Code** | **Description** |
| --- | --- |
| O00% | Ectopic pregnancy |
| O01% | Hydatidiform mole |
| O02% | Other abnormal products of conception |
| O03% | Spontaneous abortion |
| O04% | Medical abortion |
| O05% | Other abortion |
| O06% | Unspecified abortion |
| O08% | Complications following abortion and ectopic and molar pregnancy |
| O60% | Preterm labour and delivery |
| O61% | Failed induction of labour |
| O62% | Abnormalities of forces of labour |
| O63% | Long labour |
| O64% | Obstructed labour due to malposition and malpresentation of fetus |
| O65% | Obstructed labour due to maternal pelvic abnormality |
| O66% | Other obstructed labour |
| O67% | Labour and delivery complicated by intrapartum haemorrhage, not elsewhere classified |
| O68% | Labour and delivery complicated by fetal stress [distress] |
| O69% | Labour and delivery complicated by umbilical cord complications |
| O70% | Perineal laceration during delivery |
| O71% | Other obstetric trauma |
| O72% | Post-partum haemorrhage |
| O73% | Retained placenta and membranes, without haemorrhage |
| O74% | Complications of anaesthesia during labour and delivery |
| O75% | Other complications of labour and delivery, not elsewhere classified |
| O80% | Single spontaneous delivery |
| O81% | Single delivery by forceps and vacuum extractor |
| O82% | Single delivery by caesarean section |
| O83% | Other assisted single delivery |
| O84% | Multiple delivery |
| O85% | Puerperal sepsis |
| O86% | Other puerperal infections |
| O87% | Venous complications and haemorrhoids in the puerperium |
| O89% | Complications of anaesthesia during the puerperium |
| O90% | Complications of the puerperium, not elsewhere classified |
| Z37% | Outcome of delivery |
| Z38% | Liveborn infants according to place of birth and type of delivery |

**Table S7.** OPCS-4 code list for the end of pregnancy outcomes

| Code | Description |
| --- | --- |
| Q091 | Open removal of products of conception from uterus |
| Q101 | Dilation of cervix uteri and curettage of products of conception from uterus |
| Q102 | Curettage of products of conception from uterus NEC |
| Q111 | Vacuum aspiration of products of conception from uterus NEC |
| Q112 | Dilation of cervix uteri and evacuation of products of conception from uterus NEC |
| Q113 | Evacuation of products of conception from uterus NEC |
| Q114 | Extraction of menses |
| Q115 | Vacuum aspiration of products of conception from uterus using rigid cannula |
| Q116 | Vacuum aspiration of products of conception from uterus using flexible cannula |
| Q311 | Removal of products of conception from fallopian tube |
| Q58% | Delivery of terminated fetus |
| R062 | Feticide NEC |
| R14% | Surgical induction of labour |
| R15% | Other induction of labour |
| R17% | Elective caesarean delivery |
| R18% | Other caesarean delivery |
| R19% | Breech extraction delivery |
| R20% | Other breech delivery |
| R21% | Forceps cephalic delivery |
| R22% | Vacuum delivery |
| R23% | Cephalic vaginal delivery with abnormal presentation of head at delivery without instrument |
| R24% | Normal delivery |
| R25% | Other methods of delivery |
| R27% | Other operations to facilitate delivery |
| R28% | Instrumental removal of products of conception from delivered uterus |
| R29% | Manual removal of products of conception from delivered uterus |
| R30% | Other operations on delivered uterus |
| R32% | Repair of obstetric laceration |

**Figure S1.**
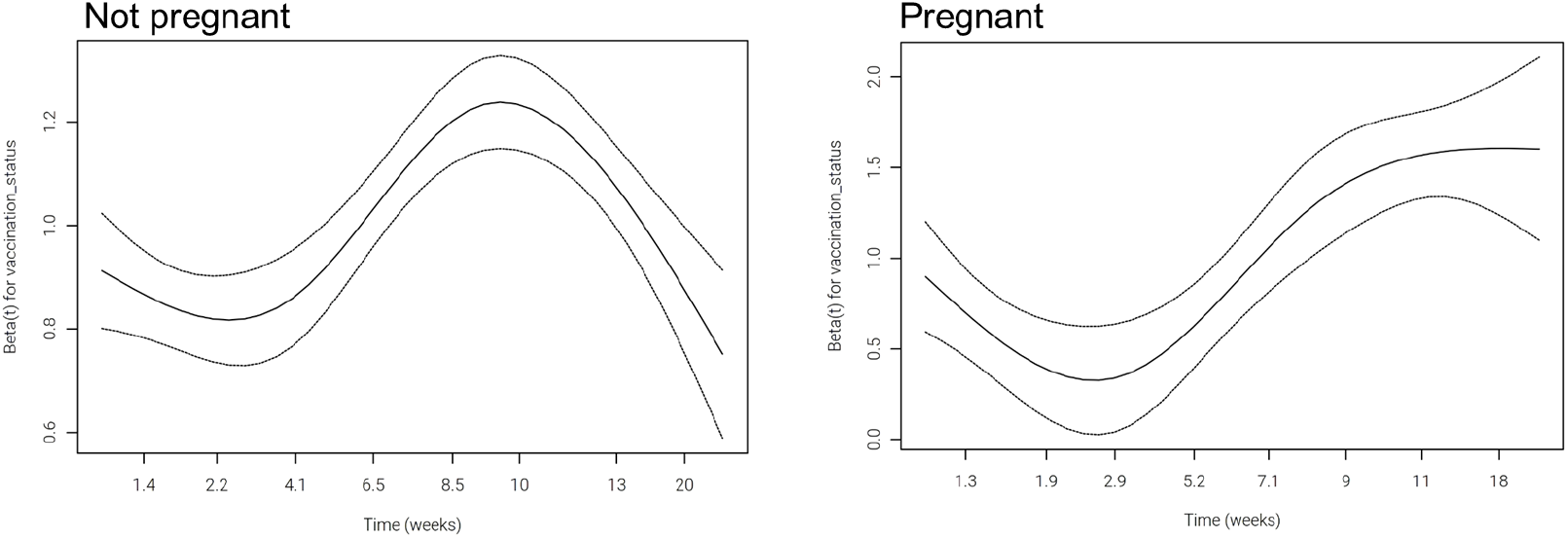
Schoenfeld residual plots for vaccination coefficient in non-pregnant (left) and pregnant (right) women.

